# Prediction of incident inflammatory bowel disease from routine electronic health records: development and external validation of a longitudinal machine learning model

**DOI:** 10.64898/2026.10.12.26365307

**Authors:** Goktug Onal, Aryan Ayati, Mao-Yuan Chen, Anshu Mukherjee, Vivek A. Rudrapatna

## Abstract

**BACKGROUND & AIMS:** Inflammatory bowel disease (IBD) is often diagnosed months to years after symptoms begin, and diagnostic delay is associated with complications. We tested whether longitudinal machine learning using electronic health record (EHR) data can identify incident IBD before diagnosis, and quantified lead time and deployment burden.

**METHODS:** We studied adults with an IBD-compatible symptom, sign, or finding at UCSF (2012-2024). Incident cases were confirmed by structured filtering and validated large language model note review. We trained a state space model (Mamba) and comparators on pre-diagnostic EHR data, excluding diagnostic-workup features, and assessed discrimination at 6- and 12-month landmarks. We externally tested Mamba without recalibration at ZSFG, a safety-net hospital serving a demographically and socioeconomically distinct population, and simulated diagnosis-code-only deployment at UCSF.

**RESULTS:** Six months after presentation, among patients still undiagnosed (67% of UCSF test cases), Mamba achieved an AUROC of 0.969 (95% CI, 0.928-0.999) at UCSF and 0.965 (0.952-0.976) at ZSFG. Median lead time was 20 months at UCSF and 15 months at ZSFG. Among future cases with an initially normal colonoscopy (N=32) or a histological finding of acute inflammation without chronicity (N=63), the model flagged 84% and 89% of these patients, respectively, before those procedures, indicating the potential for pre-diagnostic detection. Among 358,886 symptomatic patients not used for training (60 incident cases), the diagnosis-code-only model achieved an AUROC of 0.926; reviewing 20 alerts/month identified 34 cases (PPV, 1.09%; 64-fold above baseline).

**CONCLUSIONS:** Longitudinal machine learning can identify increased IBD risk months to years before diagnosis and may define a population for prospective studies of early IBD biology. Prospective evaluation should determine whether earlier identification reduces diagnostic delay.

**What You Need to Know:** *Background and Context:* IBD is often diagnosed months to years after symptoms begin. Whether routinely collected electronic health record data can identify future IBD before diagnosis, and at what alert burden, is unknown.

*New Findings:* A longitudinal model identified future IBD six months after first symptomatic presentation with an AUROC of 0.97 and a median lead time of 20 months, with similar external discrimination.

*Limitations:* This was a retrospective study; external case labels were inferred rather than adjudicated, and colonoscopy analyses were exploratory. Whether earlier flagging shortens time to diagnosis requires prospective evaluation.

*Clinical Research Relevance:* Routinely collected structured EHR data could support targeted gastroenterology review of recurrently symptomatic patients. Because incident IBD is rare among symptomatic patients, a service reviewing 20 flagged charts per month would identify roughly one future case every five months. Deployment should therefore begin in silent mode, with locally calibrated thresholds and prospective measurement of alert burden.

*Basic Research Relevance:* The model flagged most future cases before a pre-diagnostic colonoscopy read as normal, or before biopsies showing only acute inflammation. Patients flagged despite normal-appearing mucosa may represent a preclinical population in which the earliest immunologic, serologic, and microbial events of IBD could be studied prospectively, including through model-guided mucosal sampling.

*Lay Summary:* An AI system reviewing medical records data is able to identify people who would later be diagnosed with inflammatory bowel disease, often more than a year before doctors made the diagnosis.

## Introduction

Inflammatory bowel disease (IBD) often begins with nonspecific gastrointestinal symptoms — abdominal pain, altered bowel habits, rectal bleeding, fatigue — that resemble benign conditions that are more common, such as irritable bowel syndrome, hemorrhoids, and functional diarrhea.^1,2^ Recognition is therefore frequently delayed: the median interval from first symptom to diagnosis is measured in months, but a substantial minority of patients wait years.^1,3^ These delays have consequences. A longer time to diagnosis is associated with greater disease severity at presentation, a higher burden of complications, and an increased likelihood of surgery^4,5^, particularly in patients with Crohn’s disease who develop complications.^4,6^ Diagnostic delay is thus a potentially modifiable contributor to IBD morbidity.

Efforts to shorten it — red-flag symptom-awareness campaigns, fecal calprotectin testing in primary care, and streamlined referral pathways — have narrowed but not closed the gap, and delay persists even in well-resourced health systems.^7–9^ At the same time, accumulating evidence for a preclinical phase of IBD, in which systemic and mucosal inflammation are detectable before diagnostic criteria are met, indicates that a biological signal for earlier detection exists well before diagnosis is made.^10–13^ Whether that signal is already latent in the data that health systems collect in the course of routine care is unknown.

Routinely collected electronic health record (EHR) data — coded symptoms, laboratory results, medications, and patterns of health-care utilization — may encode early signatures of undiagnosed IBD.^14,15^ Prior work has achieved moderate discrimination (for example, population-based risk models with C-statistics of approximately 0.78) but shares limitations that have kept it from clinical relevance.^16^ Most approaches use static or cross-sectional feature representations that discard the temporal ordering of a patient’s encounters, even though it is the evolution of symptoms and findings over time that a clinician actually reasons over.^17,18^ Few report lead time — how far in advance of diagnosis a model generates its first alert — which is the quantity that determines whether earlier intervention is possible at all. Fewer still characterize the alert burden that deployment would impose on a health system, leaving real-world usability unquantified. And prior models have not used their own predictions to define a candidate preclinical population for prospective biological study.

We developed and evaluated a longitudinal machine learning model to predict incident IBD from structured EHR data, using a state-space sequence model (Mamba) that reads the ordered history of a patient’s encounters rather than a static summary of them. In a retrospective cohort assembled at the University of California, San Francisco (UCSF), and an external safety-net cohort, we address four questions: how well the model discriminates future cases at fixed landmarks after first symptomatic presentation, and with what lead time; whether that performance transports to a demographically and socioeconomically distinct health system without recalibration; what operating characteristics — and what alert burden — would govern deployment across the full symptomatic population; and whether patients flagged before endoscopic or histologic evidence of chronic disease constitute a population in which the earliest biology of IBD might be studied prospectively.

## Methods

### Study design and cohorts

This was a retrospective development and external-validation study of a clinical prediction model, reported in accordance with the TRIPOD+AI^19^ statement and the relevant elements of STROBE^20^ and RECORD,^21^ and approved by the UCSF Institutional Review Board with a waiver of informed consent for analysis of de-identified data. The model was developed at UCSF, an academic tertiary-care center (2012–2024), and externally validated at Zuckerberg San Francisco General Hospital (ZSFG), a safety-net hospital serving a demographically and socioeconomically distinct population (2019–2024; External validation, below). Adults (≥18 years) were eligible if they had at least one EHR-recorded symptom, sign, or finding compatible with IBD (code set, Supplementary Table S1) and at least one encounter within the study window. Cohort entry (time zero) was the date of the first qualifying finding.

We excluded patients with evidence of IBD before cohort entry — an IBD code before 2012 (predating full EHR implementation) or a documented prior diagnosis elsewhere — those whose first IBD diagnosis preceded their first qualifying finding, and those without pre-diagnostic encounters, none of which permits assessment of a pre-diagnostic window.

### Outcome ascertainment

The outcome was incident IBD during follow-up, dated at the first IBD diagnosis code (ICD-9 555–556; ICD-10 K50–K51). Because administrative codes conflate incident with established disease,^22^ cases were confirmed in two stages. First, patients with at least one IBD code were required to have documentation consistent with an IBD work-up (a gastroenterology encounter and a colonoscopy). Second, de-identified gastroenterology, colonoscopy, pathology, and discharge notes were reviewed by a large language model (GPT-4o) to confirm IBD and to exclude patients first diagnosed outside UCSF. These labels were validated against blinded manual chart review in 100 randomly selected patients (accuracy 0.96, F1 0.92; prompts and validation appear in Supplementary Methods 1.1). Controls were symptom-eligible patients never assigned an IBD code, meeting identical criteria and sampled at a 10:1 control-to-case ratio to address class imbalance.

### External validation

We externally validated the model at ZSFG using the same symptom-based eligibility criteria, exclusions, and 10:1 control-to-case ratio as at UCSF, and applied it without recalibration or threshold tuning. Because clinical notes were unavailable at ZSFG, incident status could not be adjudicated as at UCSF and was instead inferred using a structured-data classifier trained on the UCSF cohort and applied without modification (Supplementary Methods 1.2). This computable-phenotype approach substitutes an algorithmic label for chart-reviewed adjudication.^23^

### Predictors

Model inputs were structured EHR data only; clinical notes were used solely for cohort curation and were never model inputs. We harmonized ICD-9/10, CPT, and LOINC codes across demographics, diagnoses, medications, laboratory results, procedures, imaging, and encounter departments, yielding 7244 candidate features (Supplementary Table S2). To prevent leakage of the diagnostic work-up itself, features reflecting IBD evaluation or treatment — diagnostic procedures, fecal calprotectin, and IBD-specific medications and codes — were removed regardless of predictive value. Sex, race, and ethnicity were excluded from model inputs and retained only as metadata for subgroup analysis. All feature selection was performed on the training set only: features present in fewer than 1% of patients were dropped, and LASSO regression^24^ retained 204 predictors (Supplementary Table S3).

### Model development and evaluation

We trained four models spanning tabular and sequential approaches — logistic regression, gradient-boosted trees (CatBoost),^25^ a Transformer,^26^ and Mamba, a state-space sequence model that reads the ordered sequence of a patient’s encounters rather than a static summary of them^27^ (architectures and hyperparameters, Supplementary Table S4). Mamba was designated the primary model on the basis of validation-set performance, with the others reported as comparators; a variant restricted to diagnosis (ICD) codes was developed to assess portability to systems lacking richer structured data and was used for the deployment simulation. Data were split at the patient level (80% training, 10% validation, 10% test; Supplementary Table S5), stratified on case status, with every patient and all associated encounters confined to a single partition. Predictions were generated at each encounter, but all metrics were computed at the patient level, each patient contributing a single prediction and counted positive if the model flagged IBD at any pre-diagnostic encounter. Discrimination was quantified by the area under the receiver-operating-characteristic curve (AUROC) and the area under the precision–recall curve (AUPRC); calibration by the Brier score and calibration curves. Probabilities were recalibrated by isotonic regression fitted on the validation set, and operating thresholds were set on the validation set by Youden’s J^28^ and applied unchanged to both test cohorts. Discrimination and calibration metrics are reported with bootstrap 95% confidence intervals.

### Landmark discrimination and lead time

To assess whether the model could discriminate future IBD at clinically meaningful time points, we performed landmark analyses at 6 and 12 months after cohort entry.^29^ At each landmark, we restricted the analysis to patients who remained undiagnosed and under follow-up and used only prediction scores available on or before that time to predict subsequent IBD diagnosis. We reported the number of patients evaluable at each landmark. For each true-positive case, the flag date was the earliest encounter at which the prediction score exceeded the Youden’s J threshold, and lead time was the interval from flag to recorded diagnosis. An event-anchored sensitivity analysis excluded predictions occurring within 6 or 12 months of diagnosis (Supplementary Methods 1.3).

### Deployment simulation

To estimate operating characteristics under real-world use, we simulated prospective monthly deployment of the diagnosis-code model across the full symptomatic UCSF population excluding the training set (358,886 patients; 60 incident cases, corresponding to the validation- and test-set cases, because training-set patients were excluded; prevalence, 0.017%) over 2012–2024, applied without refitting. At each monthly evaluation, patients with at least one encounter in the preceding 12 months were ranked by their highest prediction score to date, the alert threshold was set to meet a fixed monthly review capacity (10, 20, 50, 100, or 200 alerts), and each patient could be alerted at most once. For each capacity we report cases identified, sensitivity, PPV, and number needed to flag (1/PPV), with Clopper– Pearson exact binomial 95% confidence intervals.^30^ Sensitivity analyses varied the evaluation cadence and scoring rule.

### Temporal robustness

To assess stability over the study period, we grouped patients into four multi-year bins by index year (year of diagnosis for cases and of last encounter for controls) and computed AUROC and AUPRC within each bin. To test for temporal drift in the model’s discriminative relationship, we fitted a logistic regression of case status on calendar year, prediction score, and their interaction; a non-significant year-by-score interaction indicates that the score’s association with the outcome did not change materially over time.

### Post-hoc and subgroup analyses

The following analyses did not influence model development. We identified pre-diagnostic IBD-related hospitalizations and compared each patient’s flag date with the date of first such admission. A validated large language model pipeline (GPT-5) classified pre-diagnostic colonoscopy and biopsy reports as showing no, acute, chronic, or mixed inflammation and compared flag dates with procedure dates (Supplementary Methods 1.4); the pipeline was validated against blinded manual review in 50 patients. Finally, at ZSFG we stratified discrimination and calibration by sex, race, ethnicity, and age band and assessed positive-prediction-rate and true-positive-rate parity at the operating threshold.

## Results

### Cohort characteristics

Of 1365 UCSF patients meeting structured inclusion criteria, large language model adjudication confirmed 410 as incident IBD diagnosed at UCSF (agreement with blinded manual review: accuracy 0.96, F1 0.92; Supplementary Table S6); 298 had at least one documented pre-diagnostic symptom or extraintestinal manifestation (see Supplementary Table S1 for the symptoms and extraintestinal manifestations) and formed the analytic case cohort (Figure 1). Of the symptom-eligible population, 3039 patients were sampled as controls at an approximately 10:1 ratio. The external ZSFG cohort comprised 139 cases and 1398 controls (full performance metrics for the structured-data incident-status classifier are provided in Supplementary Table S7). Compared with UCSF cases, ZSFG cases were older at diagnosis (median, 54 vs 37 years), more often had ulcerative colitis (74.1% vs 54.0%), had longer recorded diagnostic delays (ulcerative colitis, 371 vs 106.5 days; Crohn’s disease, 455 vs 258 days), and more healthcare encounters (median, 38 vs 16; Table 1).

**Figure 1.**
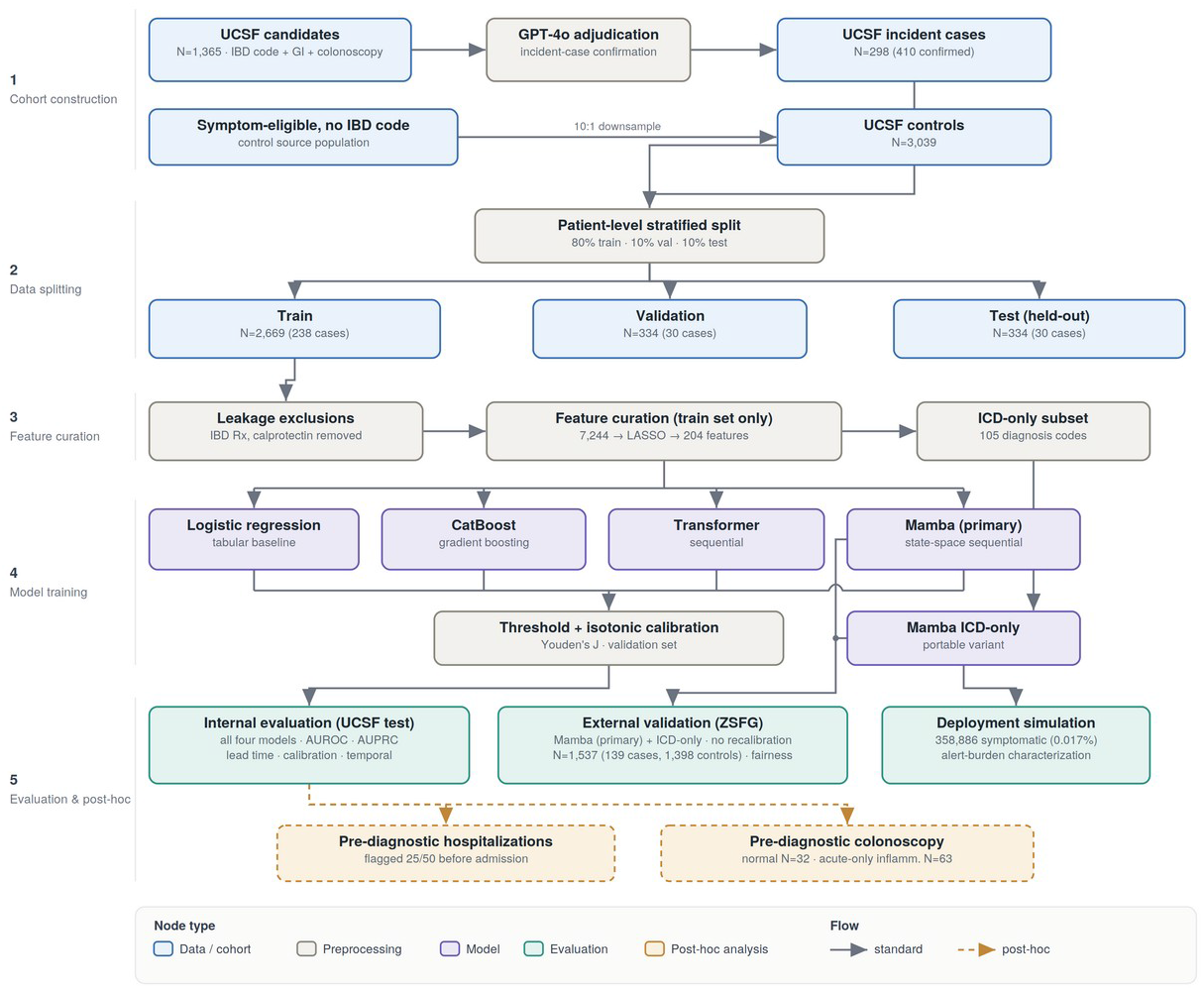
Study design and cohort derivation. Structured electronic health record (EHR) queries followed by large language model (GPT-4o) adjudication identified 410 incident inflammatory bowel disease (IBD) cases at the University of California, San Francisco (UCSF) of which 298 had at least one pre-diagnostic encounter for an IBD symptom or extraintestinal manifestation; symptom-eligible patients without an IBD code were downsampled 10:1 as controls. Data were split at the patient level into training, validation, and test partitions, and feature curation—including exclusion of diagnostic work-up signals to prevent leakage—was performed on the training set only. Four architectures and a portable diagnosis-code variant were trained, with operating-threshold selection and isotonic calibration on the validation set. Evaluation comprised the internal UCSF test set and the external Zuckerberg San Francisco General Hospital (ZSFG) cohort (primary [Mamba] and diagnosis-code models only), a deployment simulation in the full symptomatic population, and post-hoc analyses of pre-diagnostic hospitalizations and colonoscopy findings. Node colors denote node type as shown in the key. AUPRC, area under the precision–recall curve; AUROC, area under the receiver operating characteristic curve; ICD, International Classification of Diseases; LASSO, least absolute shrinkage and selection operator.

**Table 1.**
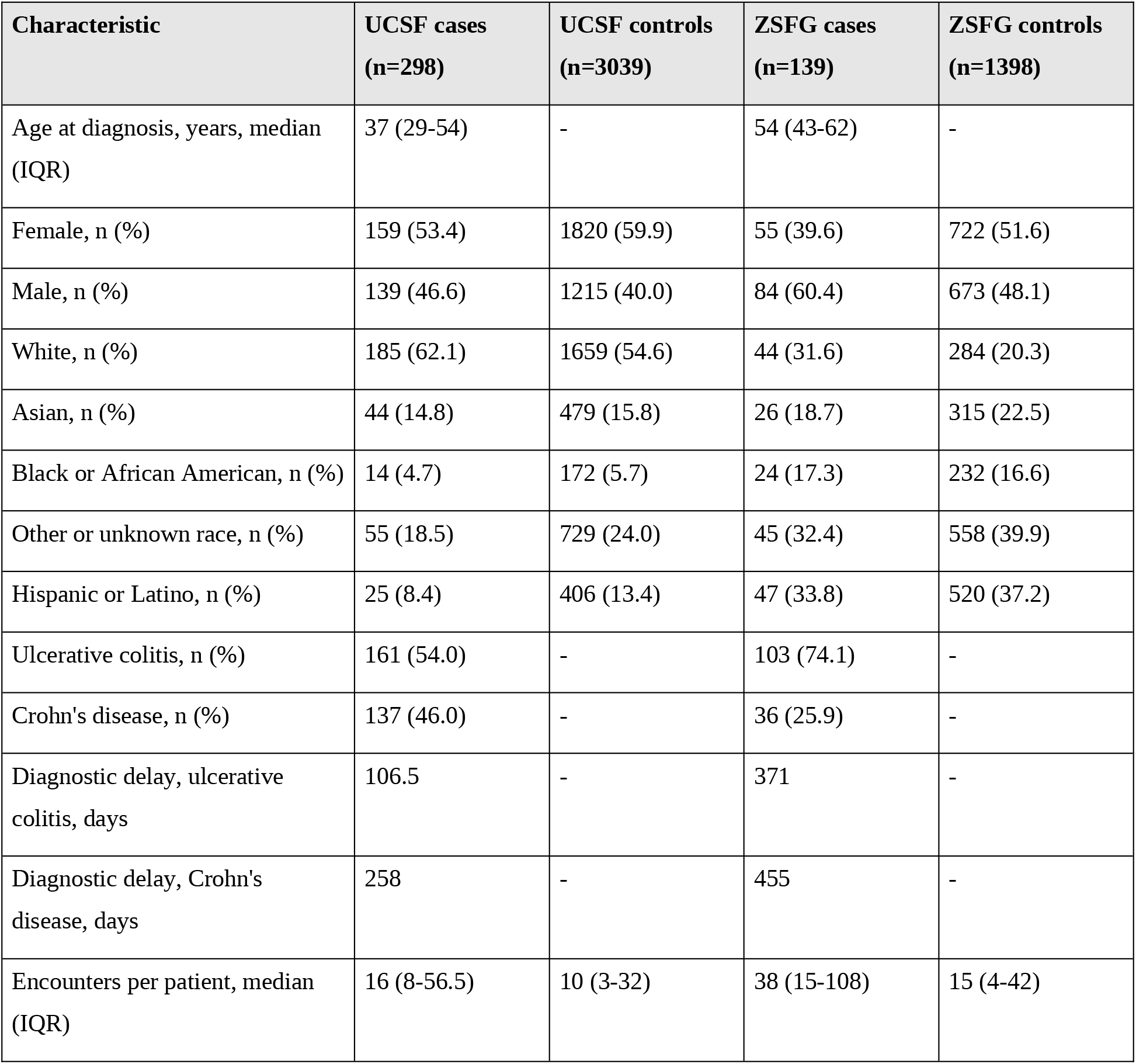
Data are median (interquartile range [IQR]) or n (%). Diagnostic delay is the median interval, in days, from first IBD-compatible presentation to recorded diagnosis; age at diagnosis and disease subtype apply to cases only. Percentages may not sum to 100 owing to rounding or missing data. IBD, inflammatory bowel disease; IQR, interquartile range; UCSF, University of California, San Francisco; ZSFG, Zuckerberg San Francisco General Hospital.

### Overall discrimination and model comparison

Across all pre-diagnostic encounters, all four models separated future IBD cases from controls (AUROC, 0.85–0.98; Table 2). Mamba had the highest AUROC (0.98; 95% CI, 0.95–1.00), followed by the Transformer (0.94; 0.89–0.98), CatBoost (0.89; 0.80–0.95), and logistic regression (0.85; 0.75–0.93). At validation-derived thresholds, Mamba also had the highest sensitivity (0.97; 0.90–1.00) and specificity (0.95; 0.92– 0.97). Because this analysis included predictions throughout the pre-diagnostic period, we used it primarily to compare model architectures; early prediction was assessed separately in the landmark analyses below.

**Table 2.** Overall rows use all pre-diagnostic encounters; landmark rows are restricted to patients still undiagnosed and under follow-up at the landmark, using only prediction scores generated on or before it. Thresholds were fixed on the UCSF validation set by Youden’s J and applied unchanged to both test cohorts. Sensitivity and specificity are independent of the 1:10 case-to-control sampling ratio; predictive values, which are not, are reported for the deployment population in Table 3. AUROC, area under the receiver operating characteristic curve; CI, confidence interval; IBD, inflammatory bowel disease; UCSF, University of California, San Francisco; ZSFG, Zuckerberg San Francisco General Hospital.

| Model | Cases | Controls | AUROC (95% CI) | Sensitivity (95% CI) | Specificity (95% CI) |
| --- | --- | --- | --- | --- | --- |
| Logistic regression (UCSF) | 30 | 304 | 0.85 (0.75-0.93) | 0.77 (0.60-0.90) | 0.85 (0.81-0.89) |
| CatBoost (UCSF) | 30 | 304 | 0.89 (0.80-0.95) | 0.77 (0.63-0.90) | 0.87 (0.83-0.90) |
| Transformer (UCSF) | 30 | 304 | 0.94 (0.89-0.98) | 0.87 (0.73-0.97) | 0.91 (0.88-0.94) |
| Mamba (UCSF) | 30 | 304 | 0.98 (0.95-1.00) | 0.97 (0.90-1.00) | 0.95 (0.92-0.97) |
| Mamba (ZSFG) | 139 | 1398 | 0.98 (0.97-0.99) | 0.94 (0.89-0.97) | 0.86 (0.84-0.88) |
| Mamba, 6-month landmark (UCSF) | 20 | 213 | 0.969 (0.928-0.999) | 0.900 (0.750-1.000) | 0.977 (0.953-0.995) |
| Mamba, 12-month landmark (UCSF) | 16 | 181 | 0.968 (0.921-1.000) | 0.875 (0.688-1.000) | 0.972 (0.945-0.994) |
| Mamba, 6-month landmark (ZSFG) | 101 | 1042 | 0.965 (0.952-0.976) | 0.703 (0.614-0.792) | 0.949 (0.936-0.962) |
| Mamba, 12-month landmark (ZSFG) | 82 | 948 | 0.967 (0.954-0.978) | 0.720 (0.622-0.805) | 0.946 (0.931-0.960) |

### Landmark discrimination

In the prespecified primary analysis — restricted at each landmark to patients still undiagnosed and under follow-up, using only scores generated on or before the landmark — Mamba discriminated future IBD with an AUROC of 0.969 (95% CI, 0.928-0.999) at 6 months and 0.968 (95% CI, 0.921-1.000) at 12 months in the UCSF test set (Table 2). Among cases evaluable at the 6-month landmark, the median interval from landmark to diagnosis was 2.4 years, indicating discrimination was not driven by patients approaching imminent diagnosis. Of 30 cases, 20 (67%) were evaluable at 6 months and 16 (53%) at 12 months; evaluability and discrimination are distinct constraints, since a patient whose entire pre-diagnostic history falls within the landmark window cannot be flagged early. In an event-anchored sensitivity analysis that counts non-evaluable patients as undetected, the 6-month AUROC was 0.67 (95% CI, 0.53-0.80) (Supplementary Table S8).

### Lead time

Median lead time was 20 months (IQR, 2-40) for Mamba and 18 months (1-46) for the Transformer, versus 4 months for both CatBoost (1-10) and logistic regression (1-11) (Figure 2). Among true-positive cases at UCSF, 55% were flagged more than one year before diagnosis, with smaller fractions flagged at 6-12 months (7%), 3-6 months (14%), 1-3 months (14%), or less than 1 month (10%) beforehand (Supplementary Table S9). Representative pre-diagnostic trajectories are shown in Figure 3.

**Figure 2.**
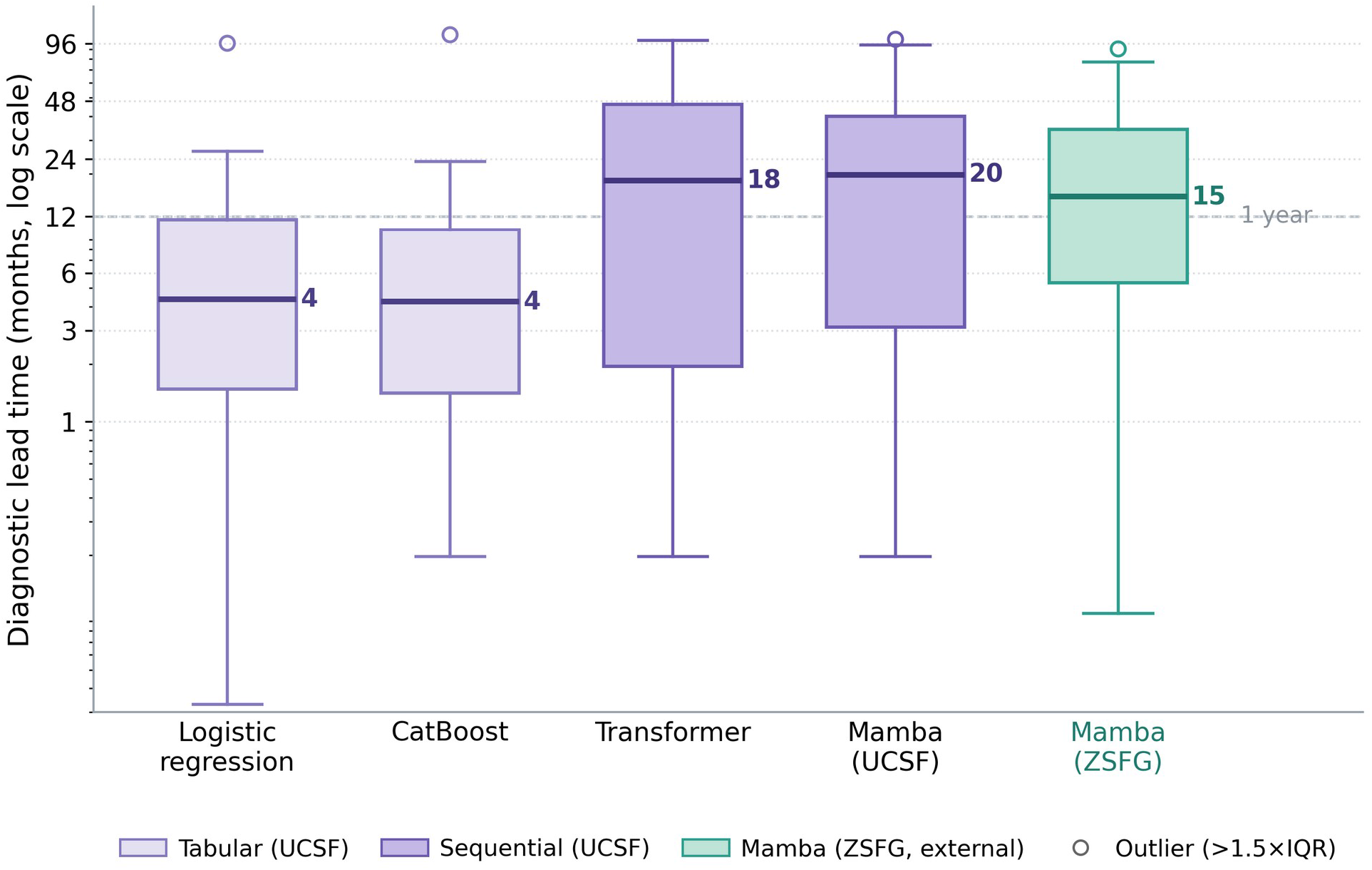
Distribution of diagnostic lead time by model. Lead time between the first model-generated flag and the recorded IBD diagnosis date among true-positive patients, in months (logarithmic scale). The center line indicates the median, boxes the interquartile range (IQR), and whiskers extend to 1.5 times the IQR; open circles denote values beyond that range. Results are shown for logistic regression, CatBoost, the Transformer, and Mamba at UCSF, and for Mamba at ZSFG. IBD, inflammatory bowel disease.

**Figure 3.**
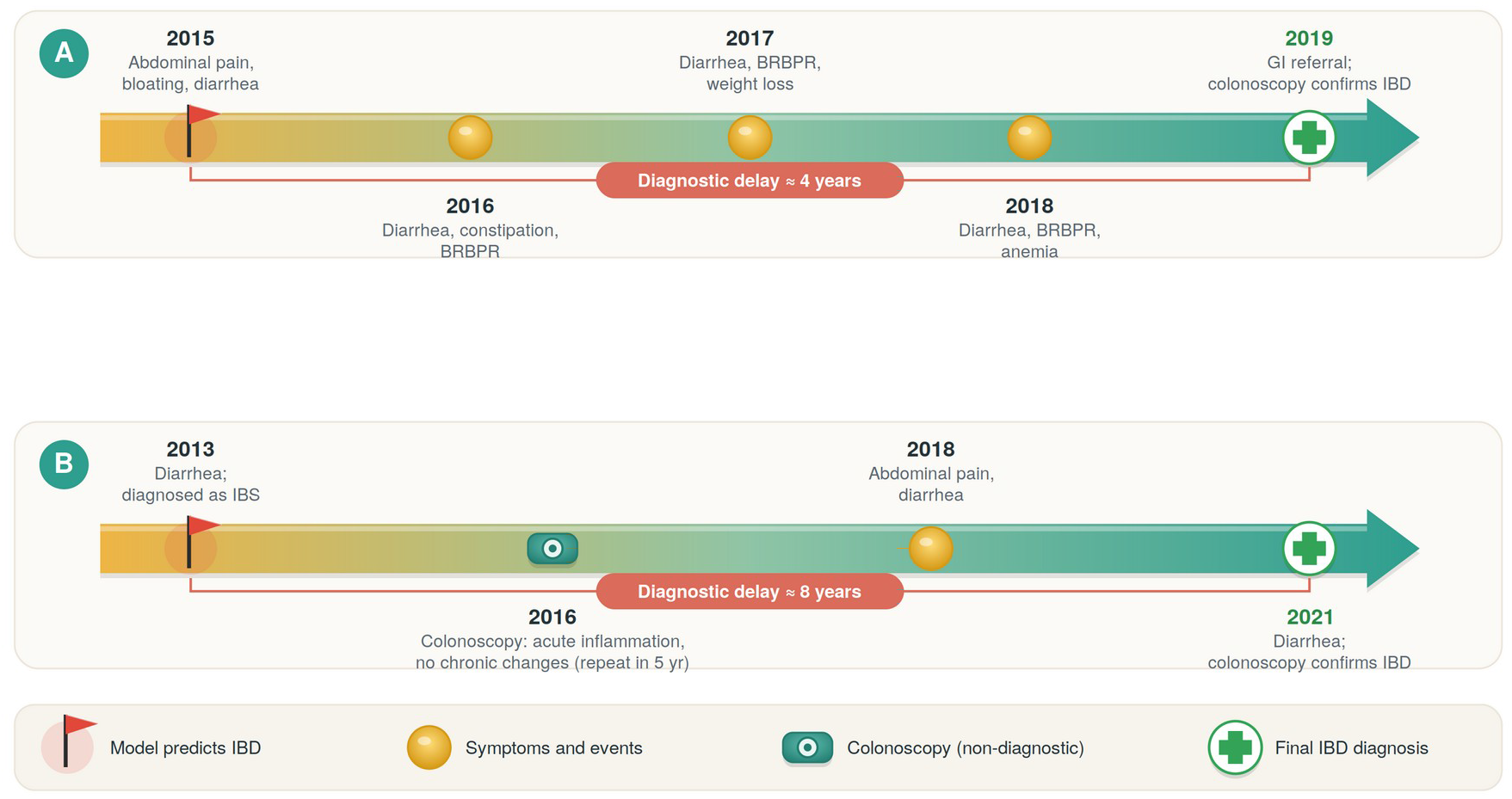
Example patient trajectories. Two illustrative patient timelines (A, B) showing model predictions in relation to clinical events. In A, symptoms accrued over approximately 4 years before colonoscopy confirmed IBD; in B, an approximately 8-year interval included a non-diagnostic colonoscopy showing acute inflammation without chronic features. Red flags indicate the first model-generated alert; yellow circles, symptom-related encounters; the endoscope symbol, a non-diagnostic colonoscopy; and green crosses, the date of confirmed IBD diagnosis. Brackets indicate the interval from first flag to diagnosis. These are illustrative cases selected to show plausible pathways to delay and are not representative of the cohort. BRBPR, bright red blood per rectum; IBS, irritable bowel syndrome.

### Calibration

After isotonic recalibration fitted on the validation set, Mamba was well calibrated in the internal test set, with a Brier score of 0.02 and predicted probabilities tracking observed frequencies (Supplementary Figure S1). At ZSFG, where no recalibration was performed, the Brier score was 0.08, similar to that of a non-informative model at the observed prevalence (0.08), indicating that recalibration would be required before local use. Calibration is reported at the 1:10 case-to-control sampling ratio and will differ at deployment prevalence.

### External validation

Applied at ZSFG without recalibration, Mamba achieved landmark AUROCs of 0.965 (95% CI, 0.952-0.976) at 6 months and 0.967 (95% CI, 0.954-0.978) at 12 months, an overall AUROC of 0.98 (95% CI, 0.97-0.99), an external AUPRC of 0.85 (95% CI, 0.78–0.91), and a median lead time of 15 months (IQR, 5-34). At the fixed UCSF threshold, sensitivity was 0.94 and specificity 0.86 (overall accuracy, 0.87; see Supplementary Table S10 for full performance metrics).

### Portable diagnosis-code model

A model restricted to diagnosis codes retained an AUROC of 0.91 (95% CI, 0.84–0.97) at UCSF and 0.95 (95% CI, 0.93–0.96) at ZSFG, with median lead times of 18 and 11 months. At the UCSF-derived threshold, sensitivity was 0.83 at UCSF and 0.66 at ZSFG, and specificity was 0.91 and 0.96 (see Supplementary Table S11 for full performance metrics). Because diagnosis codes are widely available across health systems, we used this model for the deployment simulation.

### Simulated deployment and alert burden

In simulated deployment across 358,886 symptom-eligible patients not used for model fitting (60 incident cases; prevalence, 0.017%), the diagnosis-code model retained an AUROC of 0.926. At a review capacity of 20 alerts per month, 34 of 60 cases (56.7%) were identified with a PPV of 1.09%, approximately 64-fold higher than baseline prevalence and corresponding to 92 patients reviewed per case identified. At 50 alerts per month, 44 of 60 cases (73.3%) were identified with a PPV of 0.56%, approximately 33-fold higher than baseline prevalence and corresponding to 177 patients reviewed per case identified. Increasing review capacity from 10 to 200 alerts per month increased sensitivity from 35.0% to 83.3% while PPV decreased from 1.35% to 0.16% (Table 3, Figure 4). Results were similar across reassessment intervals of 1 to 6 months (see Supplementary Table S12 for full cadence, budget and scoring rule evaluation).

**Figure 4.**
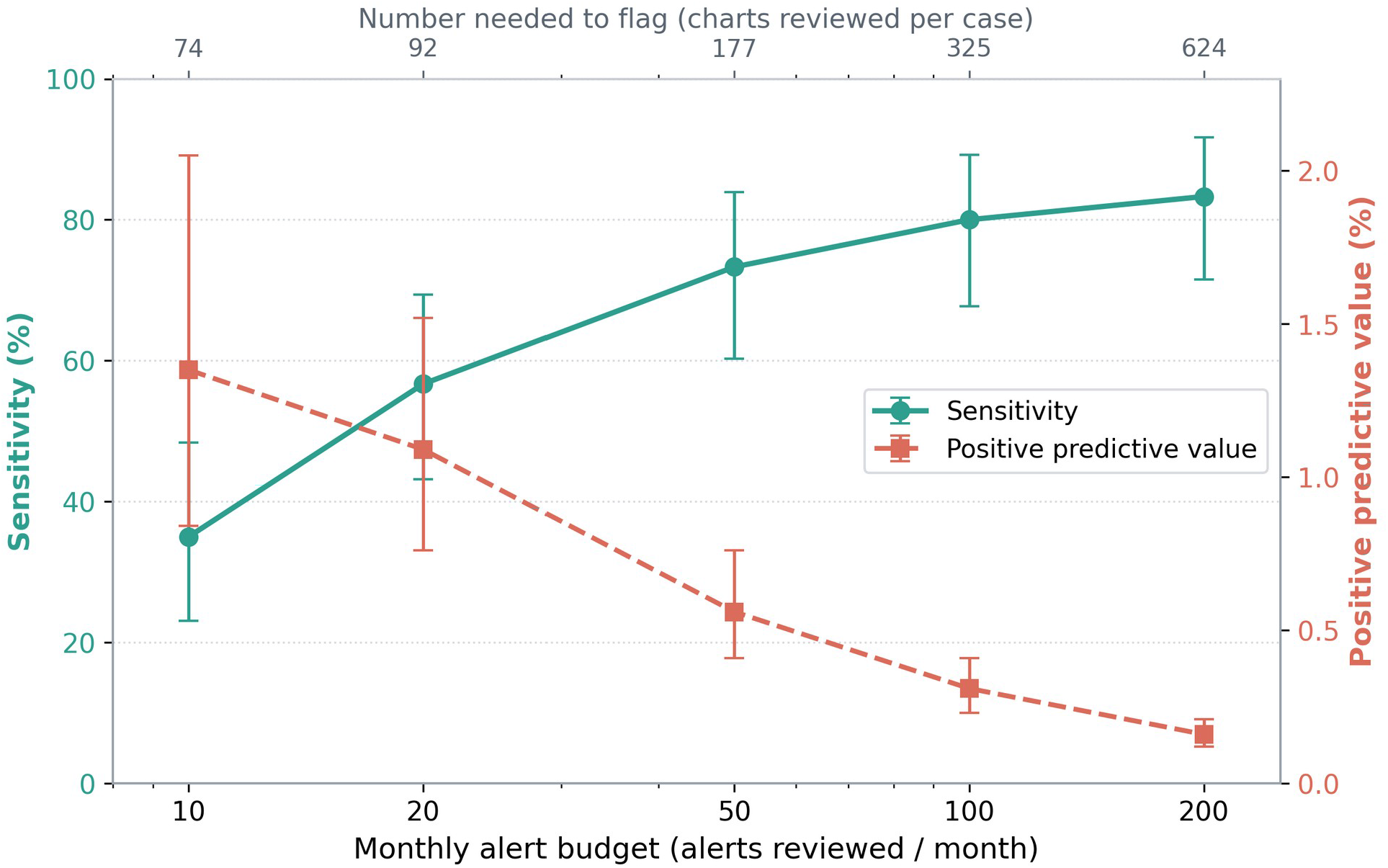
Operating characteristics of the diagnosis-code model in the full symptomatic population. Sensitivity (left axis) and positive predictive value (PPV; right axis) as functions of the monthly alert budget (10–200 alerts per month) under simulated prospective monthly deployment across 358,886 symptom-eligible patients (incident IBD prevalence, 0.017%). The corresponding number needed to flag (1/PPV; charts reviewed per case identified) is shown on the upper axis. Error bars show Clopper–Pearson exact binomial 95% confidence intervals.

**Table 3.** Diagnosis-code model applied without refitting to 358,886 symptomatic patients at UCSF (2012–2024) excluding the training set, among whom 60 developed incident IBD (prevalence 0.017%). A patient was flagged when the model score at any encounter up to the evaluation date exceeded the operating threshold, which was set at each monthly evaluation to meet the stated alert budget; each patient could be alerted at most once. Alerts issued is the total over the 156-month deployment window. Sensitivity is calculated against all 60 incident cases; 95% confidence intervals (CIs) for sensitivity and PPV are Clopper–Pearson exact binomial. Number needed to flag = 1/PPV; fold enrichment = PPV ÷ baseline incident prevalence (0.017%). CI, confidence interval; IBD, inflammatory bowel disease; PPV, positive predictive value.

| Alert budget per month | Alerts issued | Sensitivity | Specificity | PPV | Number needed to flag | True cases identified per month | Fold enrichment |
| --- | --- | --- | --- | --- | --- | --- | --- |
| 10 | 1560 | 35.0% (23.1-48.4) | 99.6% | 1.35% (0.84-2.05) | 74 | 0.13 | 79x |
| 20 | 3120 | 56.7% (43.2-69.4) | 99.1% | 1.09% (0.76-1.52) | 92 | 0.22 | 64x |
| 50 | 7800 | 73.3% (60.3-83.9) | 97.8% | 0.56% (0.41-0.76) | 177 | 0.28 | 33x |
| 100 | 15600 | 80.0% (67.7-89.2) | 95.7% | 0.31% (0.23-0.41) | 325 | 0.31 | 18x |
| 200 | 31200 | 83.3% (71.5-91.7) | 91.3% | 0.16% (0.12-0.21) | 624 | 0.32 | 9x |

### Temporal robustness

Discrimination was stable across index years 2012–2024, with AUROC at or above 0.92 and AUPRC at or above 0.85 across four multi-year bins (Supplementary Figure S2). In a test for temporal drift, the year-by-score interaction was non-significant (p = 0.50), giving no evidence that the model’s discriminative relationship changed over the study period.

### Pre-diagnostic hospitalizations

Among 50 true-positive patients with at least one pre-diagnostic IBD-related hospitalization, the model had flagged 25 (50%) before the date of their first such admission, spanning 39 pre-diagnostic IBD-related hospitalizations (mean, 1.6 per patient); the most frequent admitting diagnoses were gastrointestinal bleeding, bowel obstruction, abdominal or perirectal abscess, and severe colitis.

### Pre-diagnostic colonoscopy findings

In exploratory analyses of small subgroups, the model had flagged 27 of 32 patients (84%) before a pre-diagnostic colonoscopy reported as normal, and 56 of 63 (89%) before a pre-diagnostic colonoscopy or biopsy showing acute inflammation without chronic features — that is, before strong endoscopic or histologic evidence of chronic IBD was present. Colonoscopy and biopsy reports were categorized by a validated large language model pipeline (GPT-5; accuracy >90% against manual review in 50 patients; Supplementary Table S13).

### Fairness and subgroup performance

At ZSFG, discrimination was consistent across subgroups defined by sex, race, ethnicity, and age (AUROC range, 0.968-0.991), with comparable Brier scores; positive-prediction-rate and true-positive-rate parity were examined at the operating threshold, and across-group differences in positive predictive value tracked underlying prevalence rather than a change in model behavior (Supplementary Table S14).

## Discussion

Structured EHR data collected in routine care contain signals that identify patients who will be diagnosed with IBD months to years before the diagnosis is recorded. At a landmark 6 months after the first IBD-compatible presentation — restricted to patients still undiagnosed and using only data available at that point — a state-space sequence model discriminated future cases with an AUROC of 0.969 at UCSF and 0.965 at an independent safety-net health system to which it was transferred without recalibration, with a median lead time between first flag and recorded diagnosis of 20 and 15 months, respectively. A model using only diagnosis codes retained most of this performance and would be portable to systems lacking richer structured data. Because ZSFG cases differed from UCSF cases in age, disease subtype, recorded diagnostic delay, and healthcare utilization, the external cohort provided a stringent test of transportability, although its case labels were inferred.

Two features of these results are, in our view, more informative than the discrimination estimates themselves. First, only two-thirds of eventual cases were evaluable at the 6-month landmark; the remainder had no qualifying encounter early enough to be flagged. Discrimination among evaluable patients and the fraction of patients who are evaluable are distinct constraints, and the latter cannot be improved by better modeling — it is a property of how early patients enter care, and it bounds the early sensitivity any such system can achieve. Second, discrimination transported across sites but the operating point did not: applying the same numerical threshold to the diagnosis-code model at the external site shifted sensitivity by roughly 17 percentage points. Reports of “transportable” prediction models should distinguish these, since a model whose discrimination transfers may still require local threshold calibration before use.

Most prior machine-learning work in IBD has addressed prognosis or treatment response after diagnosis rather than earlier recognition of disease onset.^17,31^ Studies of pre-diagnostic identification have generally relied on cross-sectional or static representations in primary-care or registry cohorts, achieving moderate discrimination (AUROC ∼0.70–0.78) with limited evaluation of lead time.^16–18^ This study forecasts future diagnosis from longitudinal encounter histories, reports lead time and evaluability explicitly, validates across institutions without recalibration, and provides a portable diagnosis-code comparator; we are not aware of prior models that combine these elements or that quantify the alert burden deployment would impose. Both sequential architectures flagged patients approximately a year earlier than both tabular architectures, consistent with the temporal ordering of clinical events carrying information that cumulative counts do not, although differences in architecture and feature representation preclude attributing this difference specifically to temporal ordering.

Beyond earlier recognition, the model may illuminate the earliest phase of disease. In exploratory analyses, it flagged most patients before a pre-diagnostic colonoscopy read as normal, and most of those whose biopsies showed acute inflammation without chronic features, before those procedures took place. A visually normal colonoscopy is ordinarily treated as reassuring; that flags preceded these findings suggests structured EHR data may carry pre-diagnostic signals that precede overt endoscopic or histologic change.^10,12,13^ If this holds in larger, prospectively sampled cohorts, patients flagged despite normal-appearing mucosa would constitute an informative population for study of the earliest biological events in IBD — a preclinical phase increasingly recognized in serologic and mucosal work.^11,13^ Whether model-derived risk scores could contribute to emerging definitions of a preclinical or “pre-IBD” state, analogous to prediabetes,^10,15^ will require prospective validation, including the model-guided mucosal-sampling study we propose. These subgroups were small and were not pre-specified, and the interpretation is hypothesis-generating.

The model also identified a substantial share of patients before acute presentation: half of those who experienced a pre-diagnostic IBD-related hospitalization were flagged before their first such admission, which spanned diagnoses including gastrointestinal bleeding, bowel obstruction, abscess, and severe colitis. Because diagnostic delay in IBD is associated with greater complication and hospitalization rates,^5,6^ earlier recognition of these patients is a plausible target for reducing preventable acute-care morbidity.

### Clinical implications and deployment

Potential deployment approaches include identifying recurrently symptomatic patients for targeted gastroenterology evaluation and prioritizing endoscopy among patients already referred; both require prospective evaluation. Each depends on an alert burden a service can absorb, and we therefore regard the full symptomatic-population analysis, rather than the discrimination estimates, as the result that determines whether this model is usable. In that population, a triage service reviewing 50 charts per month would identify roughly one future IBD patient every three to four months, at approximately 177 charts per case. Despite an AUROC of 0.926, precision was low in the deployment simulation (AUPRC 0.007). The low case fraction contributed to the review burden, although improvements in discrimination could increase precision. Evaluation in clinically enriched populations may therefore be useful, but their operating characteristics must be measured directly. Any deployment should begin in silent mode, generating predictions without clinical action to measure alert volume, achieved lead time, and likely response before any practice change, with thresholds calibrated locally rather than transferred, continuous monitoring for calibration drift, and predictions routed through a clinician-reviewed pathway rather than presented as interruptive alerts.

### Limitations

This study is retrospective and shows that cases can be flagged early, not that flagging shortens time to diagnosis; the latter requires prospective evaluation and, ultimately, a pragmatic trial. The external cohort is the principal source of uncertainty. Because clinical notes were unavailable at ZSFG, incident status was inferred by a structured-data classifier rather than adjudicated. At ZSFG, a substantial fraction of external cases may be prevalent rather than incident, and prevalent contamination would inflate apparent external discrimination; the short external window (2019–2024) further limits the lookback available to separate incident from established disease. Defining cases by an unvalidated, transported computable phenotype and then using those labels for evaluation departs from the usual practice of reserving adjudicated labels for performance assessment,^23^ and we therefore treat the external results as supportive rather than confirmatory. In addition, a patient was counted positive if flagged at any pre-diagnostic encounter, so patients with more encounters had more opportunities to be flagged; cases had more encounters than controls at both sites, and part of the observed discrimination may reflect healthcare-utilization intensity rather than pre-diagnostic disease. Controls were sampled at approximately 1:10, so the predictive values reported here do not correspond to deployment prevalence. In the deployment simulation, training-set patients were excluded, so incident IBD was under-represented (0.017% vs approximately 0.08% in the full symptomatic population), and predictive values there are therefore conservative. Cohort curation depended on large language model adjudication, which is sensitive to prompt design and note completeness, and some misclassification is likely despite two-stage review. Predictors were limited to structured data; clinical notes, imaging, and pathology narratives probably carry further signal. Subgroup analyses were limited to the external site, rested on inferred case labels, and were imprecise for smaller subgroups; similar performance across measured subgroups does not establish the absence of bias. Finally, both sites lie within a single metropolitan region and health-system family, and validation in other systems, EHR vendors, and countries remains necessary.

### Conclusions

Routinely collected structured EHR data contain pre-diagnostic signatures of IBD that are detectable at clinically actionable horizons and transport across two distinct health systems. Whether this capability shortens time to diagnosis depends on questions this study does not answer: how many alerts a service must absorb per case identified, whether clinicians act on those alerts, and whether earlier action improves outcomes. Prospective, silent-mode evaluation with local calibration — paired, where feasible, with model-guided sampling of patient tissue before the onset of IBD — is the necessary next step.

## Declaration of generative AI and AI-assisted technologies in manuscript preparation

During preparation of this manuscript, the authors used Claude (Anthropic) and ChatGPT (OpenAI) to assist with language editing, organization, internal consistency checks, and manuscript revision. The authors reviewed and edited all output and take full responsibility for the content of the publication.

## Author contributions (CRediT)

G.O.: Conceptualization, Methodology, Software, Formal analysis, Data curation, Visualization, Writing – original draft. A.A.: Data curation, Validation (manual chart review), Writing – review & editing. M.-Y.C.: Validation (manual chart review), Writing – review & editing. A.M.: Validation (manual chart review). V.A.R.: Conceptualization, Methodology, Supervision, Resources, Funding acquisition, Writing – review & editing. All authors approved the final manuscript.

## Ethics

The University of California, San Francisco Institutional Review Board (protocol 18-24588) approved this study, including research at Zuckerberg San Francisco General Hospital, and waived the requirement for informed consent for analysis of de-identified data.

## Data Availability Statement

The patient-level electronic health record data used in this study are subject to privacy and institutional restrictions and cannot be shared publicly. Aggregate results necessary to support the findings are provided in the manuscript and Supplementary Materials. Analysis code is available from the corresponding author upon reasonable request.

## Professional medical writing assistance

None.

## References

1. Cross E, Saunders B, Farmer AD, et al. Diagnostic delay in adult inflammatory bowel disease: A systematic review. Indian J Gastroenterol 2023;42:40–52.

2. AWARE-IBD Diagnostic Delay Working Group. Sources of diagnostic delay for people with Crohn’s disease and ulcerative colitis: Qualitative research study. PLoS ONE 2024;19:e0301672.

3. Souaid C, Fares E, Primard P, et al. A review investigating delays in Crohn’s disease diagnosis. Clinics and Research in Hepatology and Gastroenterology 2025;49:102500.

4. Ricciuto A, Mack DR, Huynh HQ, et al. Diagnostic Delay Is Associated With Complicated Disease and Growth Impairment in Paediatric Crohn’s Disease. J Crohns Colitis 2020;15:419–431.

5. Lee D, Koo JS, Choe JW, et al. Diagnostic delay in inflammatory bowel disease increases the risk of intestinal surgery. WJG 2017;23:6474–6481.

6. Schoepfer A, Santos J, Fournier N, et al. Systematic Analysis of the Impact of Diagnostic Delay on Bowel Damage in Paediatric Versus Adult Onset Crohn’s Disease. Journal of Crohn’s and Colitis 2019;13:1334–1342.

7. Perry RW, Mummery D, Alexander JL, et al. Faecal calprotectin in primary care: is its current use supported by the evidence? Br J Gen Pract 2024;74:134–136.

8. Vernon-Roberts A, Day AS. Promoting early testing and appropriate referral to reduce diagnostic delay for children with suspected inflammatory bowel disease, a narrative review. Transl Pediatr 2023;12:1416–1430.

9. IBD UK. Clear referral pathways. Available at: https://ibduk.org/ibd-standards/pre-diagnosis/referral-pathways [Accessed May 26, 2026].

10. Rodríguez-Lago I, Zabana Y, Barreiro-de Acosta M. Diagnosis and natural history of preclinical and early inflammatory bowel disease. Ann Gastroenterol 2020;33:443–452.

11. Rudbaek JJ, Agrawal M, Torres J, et al. Deciphering the different phases of preclinical inflammatory bowel disease. Nat Rev Gastroenterol Hepatol 2024;21:86–100.

12. Bergemalm D, Andersson E, Hultdin J, et al. Systemic Inflammation in Preclinical Ulcerative Colitis. Gastroenterology 2021;161:1526–1539.e9.

13. Torres J, Petralia F, Sato T, et al. Serum Biomarkers Identify Patients Who Will Develop Inflammatory Bowel Diseases Up to 5 Years Before Diagnosis. Gastroenterology 2020;159:96–104.

14. Rajkomar A, Oren E, Chen K, et al. Scalable and accurate deep learning with electronic health records. npj Digital Med 2018;1:18.

15. Lopes EW, Turpin W, Croitoru K, et al. Prediction and Prevention of Inflammatory Bowel Disease. Clinical Gastroenterology and Hepatology 2025;23:396–405.e1.

16. Umar N, Wambua S, Harvey P, et al. Development and validation of a risk prediction tool for the diagnosis of inflammatory bowel disease in patients presenting in primary care with abdominal symptoms. J Crohns Colitis 2025;19:jjaf044.

17. Nguyen NH, Picetti D, Dulai PS, et al. Machine Learning-based Prediction Models for Diagnosis and Prognosis in Inflammatory Bowel Diseases: A Systematic Review. J Crohns Colitis 2021;16:398–413.

18. Kraszewski S, Szczurek W, Szymczak J, et al. Machine Learning Prediction Model for Inflammatory Bowel Disease Based on Laboratory Markers. Working Model in a Discovery Cohort Study. J Clin Med 2021;10:4745.

19. Collins GS, Moons KGM, Dhiman P, et al. TRIPOD+AI statement: updated guidance for reporting clinical prediction models that use regression or machine learning methods. BMJ 2024;385:e078378.

20. von Elm E, Altman DG, Egger M, et al. Strengthening the reporting of observational studies in epidemiology (STROBE) statement: guidelines for reporting observational studies. BMJ 2007;335:806–808.

21. Benchimol EI, Smeeth L, Guttmann A, et al. The REporting of studies Conducted using Observational Routinely-collected health Data (RECORD) Statement. PLoS Med 2015;12:e1001885.

22. Dawwas GK, Weiss A, Constant BD, et al. Development and Validation of Claims-Based Definitions to Identify Incident and Prevalent Inflammatory Bowel Disease in Administrative Healthcare Databases. Inflammatory Bowel Diseases 2023;29:1993–1996.

23. Pacheco JA, Rasmussen LV, Kiefer RC, et al. A case study evaluating the portability of an executable computable phenotype algorithm across multiple institutions and electronic health record environments. Journal of the American Medical Informatics Association 2018;25:1540–1546.

24. Tibshirani R. Regression Shrinkage and Selection Via the Lasso. Journal of the Royal Statistical Society Series B: Statistical Methodology 1996;58:267–288.

25. Prokhorenkova L, Gusev G, Vorobev A, et al. CatBoost: unbiased boosting with categorical features. In: Proceedings of the 32nd International Conference on Neural Information Processing Systems. NIPS’18. Red Hook, NY, USA: Curran Associates Inc.; 2018:6639–6649. Available at: https://dl.acm.org/doi/10.5555/3327757.3327770 [Accessed May 26, 2026].

26. Vaswani A, Shazeer N, Parmar N, et al. Attention is All you Need. In: Advances in Neural Information Processing Systems. Vol 30. Curran Associates, Inc.; 2017. Available at: https://proceedings.neurips.cc/paper_files/paper/2017/hash/3f5ee243547dee91fbd053c1c4a845aa-Abstract.html [Accessed May 26, 2026].

27. Gu A, Dao T. Mamba: Linear-Time Sequence Modeling with Selective State Spaces. 2024. Available at: http://arxiv.org/abs/2312.00752 [Accessed May 25, 2026].

28. Youden WJ. Index for rating diagnostic tests. Cancer 1950;3:32–35.

29. Dafni U. Landmark analysis at the 25-year landmark point. Circ Cardiovasc Qual Outcomes 2011;4:363–371. doi:10.1161/CIRCOUTCOMES.110.957951.

30. Clopper CJ, Pearson ES. The use of confidence or fiducial limits illustrated in the case of the binomial. Biometrika 1934;26:404–413. doi:10.1093/biomet/26.4.404.

31. Stafford IS, Gosink MM, Mossotto E, et al. A Systematic Review of Artificial Intelligence and Machine Learning Applications to Inflammatory Bowel Disease, with Practical Guidelines for Interpretation. Inflamm Bowel Dis 2022;28:1573–1583.

