## Supplementary Methods, Tables, and Figures for "Prediction of incident inflammatory bowel disease from routine electronic health records: development and external validation of a longitudinal machine learning model"

##### 1.1 GPT-4o adjudication of incident IBD

To establish ground truth for incident (de novo) IBD status, 100 patients were randomly selected from the 1,365 meeting initial inclusion criteria. Three reviewers (two physicians, one medical student) manually reviewed de-identified gastroenterology and colonoscopy notes; each reviewed a distinct subset, so no inter-reviewer adjudication was required. A case was incident if IBD (Crohn's disease or ulcerative colitis) was first recorded at UCSF with no evidence of prior diagnosis elsewhere. GPT-4o was applied to the same notes and its answers compared with these reference labels (validation, Table S6). The validated workflow was then applied to all 1,365 candidates to confirm incident status at scale.

**Verbatim prompt supplied to GPT-4o:**

```
I will provide you with the UCSF notes for a patient, and I would like you to answer the
following questions:
1. Does this patient have a documented diagnosis of IBD? Respond with Yes or No.
2. Is there evidence that the patient was already diagnosed with IBD at another center before
coming to UCSF? Respond with Yes or No.

At the end of your reasoning, provide your answers in the following JSON format:
{
  "ibd_diagnosis_confirmed": 0,   // 1 = Yes, 0 = No
  "diagnosed_elsewhere_before_ucsf": 0,   // 1 = Yes, 0 = No
}
Here are the patient's notes; all the notes provided are from UCSF:
```

##### 1.2 External validation cohort (ZSFG) and structured-data classifier

For external validation we identified adults (>=18 years) with >=1 IBD diagnosis code at ZSFG after 2012 and no UCSF encounters (n = 669). Because clinical notes were unavailable at ZSFG, incident status was inferred with a structured-data classifier trained at UCSF on 10,473 patients with post-2012 IBD codes (410 incident cases, 10,063 non-incident controls). Features spanned diagnoses (ICD-9 5xx, ICD-10 Kxx), IBD-related and immunomodulatory medication classes, colonoscopy counts, and Primary Care / Gastroenterology encounter indicators, plus 24 engineered temporal features (e.g., GI_Before_Colonoscopy_Count). Patients were right-truncated at their final encounter; non-engineered features were cumulatively summed and engineered features used as static counts. A CatBoost classifier with balanced class weights and a fixed 0.05 threshold was applied unchanged to the 669 ZSFG patients, classifying 139 as likely incident (classifier performance, Table S7).

#### 1.3 Event-anchored sensitivity analysis

As a lower-bound complement to the landmark analysis, an event-anchored design excluded predictions within 6 or 12 months of diagnosis and counted patients with no earlier evaluable encounter as undetected. This conflates discrimination with data availability and is reported for comparison only (results, Table S8).

#### 1.4 GPT-5 colonoscopy and biopsy annotation

**A validated GPT-5 pipeline classified pre-diagnostic colonoscopy and biopsy reports as showing no, acute, chronic, or mixed inflammation (validation, Table S13). Verbatim prompt supplied to GPT-5:**

```
You are a clinical text analyst trained to interpret medical notes.
Your task is to determine whether the text below is a colonoscopy report and, if so, to
extract structured information about inflammation.

STEP 1 - DETERMINE IF THIS IS A COLONOSCOPY REPORT
Decide whether this note documents a colonoscopy or lower endoscopy. Indicators include
explicit terms (colonoscopy, scope, endoscopy, sigmoid, rectum, cecum), descriptions of colon
segments, procedural language (scope advanced, withdrawal time, mucosal appearance), or
mentions of colon biopsies/mucosal findings. If none are present, classify as
"Is_colonoscopy_report": "No", set all other fields to "None", and skip remaining steps.

STEP 2 - IDENTIFY INFLAMMATION (ONLY IF COLONOSCOPY REPORT)
Determine whether there is any mention of inflammation in the colon, rectum, or terminal
ileum (erythema, friability, edema, granularity, loss of vascular pattern, exudate,
ulceration, mucosal breaks; colitis, proctitis, ileitis, inflammatory changes, active
inflammation). Do NOT count inflammation clearly attributed to a non-IBD cause (SCAD,
infectious colitis, ischemic colitis, radiation proctitis, NSAID colopathy, prep-related
changes, isolated inflammatory polyps without surrounding inflammation). If none:
"Inflammation_present": "No", "Type": "None", "Supporting_text": "No inflammation noted."

STEP 3 - CLASSIFY INFLAMMATION TYPE (ONLY IF INFLAMMATION PRESENT)
Acute: acute, ulceration, erosion, neutrophilic, cryptitis, crypt abscess. Chronic: chronic,
crypt distortion, basal plasmacytosis, granularity, pseudopolyps. Both: both acute and
chronic terms present. Unspecified: inflammation described but none of the above, or no
histology/pathology context. If no pathology/biopsy section and no explicit
"acute"/"chronic", classify as "Unspecified".

STEP 4 - OUTPUT FORMAT
Provide a short reasoning paragraph (1-3 sentences), then output JSON:
{
  "Is_colonoscopy_report": "Yes or No",
  "Inflammation_present": "Yes or No or None",
  "Type": "Acute / Chronic / Both / Unspecified / None",
  "Supporting_text": "Exact phrase(s) supporting the classification"
}

Now analyze this note:
```

### Supplementary Tables

#### Table S1. IBD-compatible symptom and extraintestinal-manifestation codes

| Category | ICD-10 | ICD-9 | Clinical meaning |
|---|---|---|---|
| Abdominal pain / diarrhea / rectal bleeding | R10, R19.7, K62.5 | 789, 787.91, 569.3 | Common pre-diagnostic GI symptoms |
| Fistula and fissures | K60, K62.6 | 569.41 | Perianal complications |
| Weight loss | R63.4 | 783.21 | Constitutional symptom |
| Uveitis | H20, H30 | 364, 363 | Ocular manifestation |
| Anemia | D64.9, D50 | 285.9, 280 | Systemic manifestation |
| Enteropathic arthropathy / spondyloarthropathy | M07, M45, M46 | 713.1, 720 | Joint involvement |
| Osteoporosis | M80, M81 | 733.0, 733.1 | Bone involvement |
| Erythema nodosum | L52 | 695.2 | Dermatologic manifestation |
| Pyoderma gangrenosum | L88 | 686.1 | Dermatologic manifestation |
| Primary sclerosing cholangitis | K83.01 | 576.1 | Hepatobiliary involvement |
| Autoimmune hepatitis | K75.4 | 571.49 | Hepatobiliary involvement |
| Mucus in stool / bloating / nausea / vomiting | R19.4, R14, R11, R19.5 | 787.91, 787.3, 787.0, 787.7 | GI symptoms |
| Hemorrhoids / diverticular disease | K64, K57 | 455, 562 | Mimics / confounders of IBD |

*Codes defined the pre-diagnostic prediction window for cases and controls. Percent-sign wildcards from the source are omitted for readability.*

#### Table S2. Structured feature sources in the analytic dataset

| Domain | Description | Preprocessing / harmonization | Representation |
|---|---|---|---|
| Demographics | Age at encounter | Sex, race, ethnicity excluded from inputs | Continuous (years) |
| Diagnoses | ICD-9/10 codes | Truncated to first three characters | Multi-hot per encounter |
| Medications | Inpatient/outpatient orders | Mapped to pharmaceutical subclasses | Binary per class |
| Laboratory tests | Lab results | LOINC-mapped; abnormal flagged; raw values excluded | Binary abnormal per class |
| Procedures | Surgical/diagnostic | CPT grouped by family | Binary per group |
| Imaging | Radiology orders | CPT subset | Binary per type |
| Encounter department | Department per encounter | Categorical | One-hot per department |

*7,244 unique structured features were extracted and harmonized (ICD-9/10, CPT, LOINC).*

#### Table S3. Predictors retained after LASSO regularization

| Domain | Selected (n) | Representative examples |
|---|---|---|
| Diagnoses (ICD-9/10) | 105 | K58 (IBS), K59 (constipation), R19, D64 (anemia), M54 |
| Laboratory tests (LOINC) | 11 | Hemoglobin, hematocrit, CRP, direct bilirubin, triglycerides, transferrin |
| Medication classes | 36 | Corticosteroids, SSRIs, laxatives, antibiotics, antiemetics, antifungals |
| Procedures (CPT) | 43 | Perianal abscess I&D, seton placement, MRI pelvis, abdominal ultrasound |
| Encounter departments | 6 | Primary care, GI, general internal medicine, pediatrics, immunization, rheumatology |
| Imaging | 3 | Abdominal ultrasound |
| Total | 204 | |

*Rare features (<1% prevalence) were excluded. Direct diagnostic-confirmation features (e.g., fecal calprotectin, IBD-specific medications) were removed to prevent leakage.*

#### Table S4. Model architectures and key hyperparameters

| Model | Description | Key hyperparameters |
|---|---|---|
| Logistic regression | Linear benchmark | L2 penalty (C = 1.0); solver = LBFGS |
| CatBoost | Gradient-boosted trees for sparse tabular data | Iterations = 2,000; lr = 0.008; depth = 8; loss = Logloss; early stopping = 300 rounds |
| Transformer | Visit-level temporal model | Max length = 1,024; embed dim = 128; layers = 1; heads = 2; dropout = 0.1; AdamW (lr 3e-4, wd 1e-4); batch 32; 50 epochs; BCE; early stop patience 5 (val AUPRC); 1 warmup |
| Mamba | State-space sequence model | Layers = 2; hidden 16; conv 4; expand 2; dropout 0.1; dt_rank 16; dt_scale 0.01; dt_max 0.1; max length 1,024; AdamW (lr 3e-4, wd 1e-4); batch 32; 50 epochs; BCE; early stop patience 5 (val AUPRC); 1 warmup |

*All models trained on the training set only with balanced class weights; early stopping on validation AUPRC. BCE = binary cross-entropy.*

#### Table S5. Data-split summary

| Metric | Training | Validation | Test |
|---|---|---|---|
| Patients (n) | 2,669 | 334 | 334 |
| Visits (n) | 79,880 | 12,329 | 11,233 |
| IBD cases (n, %) | 238 (8.9%) | 30 (8.9%) | 30 (8.9%) |
| Controls (n, %) | 2,431 (91.0%) | 304 (91.0%) | 304 (91.0%) |
| Case:control ratio | 1:10 | 1:10 | 1:10 |

*Patient-level stratified split (80/10/10); each patient and all encounters confined to one partition.*

#### Table S6. Validation of GPT-4o adjudication against manual review (n = 100)

| Metric | Point estimate | 95% CI (bootstrap) |
|---|---|---|
| Sensitivity | 0.923 | 0.805 - 1.000 |
| Specificity | 0.973 | 0.931 - 1.000 |
| Precision (PPV) | 0.923 | 0.805 - 1.000 |
| Accuracy | 0.960 | 0.920 - 0.990 |
| F1 score | 0.923 | 0.829 - 0.984 |

*95% CIs from non-parametric bootstrap (5,000 iterations, percentile method).*

#### Table S7. Structured-data classifier for incident IBD (UCSF test set, threshold = 0.05)

| Metric | Mean | 95% CI |
|---|---|---|
| AUROC | 0.970 | 0.944 - 0.991 |
| AUPRC | 0.758 | 0.638 - 0.870 |
| Sensitivity | 0.853 | 0.725 - 0.950 |
| Specificity | 0.942 | 0.927 - 0.956 |
| Positive predictive value | 0.380 | 0.317 - 0.452 |
| Negative predictive value | 0.994 | 0.988 - 0.998 |
| F1 score | 0.525 | 0.448 - 0.602 |

*95% CIs from non-parametric bootstrap (1,000 iterations, percentile method).*

#### Table S8. Event-anchored sensitivity analysis results, Mamba model.

| Analysis | AUROC (95% CI) | Sensitivity (95% CI) | Specificity (95% CI) | PPV (95% CI) | NPV (95% CI) |
|---|---|---|---|---|---|
| 6-month truncation (UCSF) | 0.67 (0.53-0.80) | 0.60 (0.42-0.77) | 0.95 (0.93-0.97) | 0.55 (0.38-0.71) | 0.96 (0.94-0.98) |
| 12-month truncation (UCSF) | 0.60 (0.44-0.75) | 0.53 (0.35-0.71) | 0.95 (0.93-0.97) | 0.52 (0.33-0.69) | 0.95 (0.93-0.98) |
| 6-month truncation (ZSFG) | 0.70 (0.63-0.77) | 0.68 (0.60-0.76) | 0.86 (0.84-0.88) | 0.33 (0.27-0.38) | 0.97 (0.95-0.97) |
| 12-month truncation (ZSFG) | 0.57 (0.48-0.64) | 0.51 (0.43-0.59) | 0.86 (0.84-0.88) | 0.27 (0.21-0.32) | 0.95 (0.93-0.96) |

Predictions within the specified window before diagnosis were discarded. Patients with no evaluable encounter outside that window were counted as not detected. These estimates therefore combine discrimination with data availability and are systematically lower than the landmark estimates in Table 2 of the main text; the two are not directly comparable.

#### Table S9. Lead-time distribution (Mamba, UCSF true-positive cases)

| Window before diagnosis | Definition | n | % |
|---|---|---|---|
| >12 months | >365 days prior | 16 | 55.2 |
| 6-12 months | 181-365 days prior | 2 | 6.9 |
| 3-6 months | 91-180 days prior | 4 | 13.8 |
| 1-3 months | 31-90 days prior | 4 | 13.8 |
| <1 month | 1-30 days prior | 3 | 10.3 |
| Total | - | 29 | 100.0 |

#### Table S10. Mamba external validation performance (ZSFG)

| Metric | Estimate | 95% CI |
|---|---|---|
| AUROC | 0.98 | 0.97 - 0.99 |
| AUPRC | 0.85 | 0.78 - 0.91 |
| Sensitivity | 0.94 | 0.89 - 0.97 |
| Specificity | 0.86 | 0.84 - 0.88 |
| PPV | 0.40 | 0.37 - 0.44 |
| NPV | 0.99 | 0.99 - 1.00 |
| F1 score | 0.56 | 0.53 - 0.60 |
| Brier | 0.08 | - |

*139 incident cases, 1,398 controls; UCSF-derived thresholds without recalibration. Point estimates with 95% CIs from a stratified percentile bootstrap (2,000 replicates).*

#### Table S11. Diagnosis-code (ICD-only) model performance

| Metric | UCSF (test) | ZSFG (external) |
|---|---|---|
| AUROC | 0.91 (0.84-0.97) | 0.95 (0.93-0.96) |
| AUPRC | 0.55 (0.41-0.73) | 0.60 (0.53-0.69) |
| Sensitivity | 0.83 (0.70-0.97) | 0.66 (0.58-0.74) |
| Specificity | 0.91 (0.88-0.94) | 0.96 (0.95-0.97) |
| PPV | 0.48 (0.39-0.59) | 0.63 (0.56-0.70) |
| NPV | 0.98 (0.97-1.00) | 0.97 (0.96-0.97) |
| F1 score | 0.61 (0.51-0.71) | 0.65 (0.58-0.71) |
| Brier | 0.05 | 0.06 |
| Lead time, months, median (IQR) | 18 (1-36) | 11 (4-30) |

*Thresholds set on the UCSF validation set. 95% CIs from a stratified percentile bootstrap (2,000 replicates). Feature dimensionality 204 -> 105.*

#### Table S12. Deployment simulation sensitivity analyses

| Budget | Scoring rule | Cadence (mo) | Score window | Cases (of 60) | PPV | Sensitivity |
|---|---|---|---|---|---|---|
| 10 | Running max | 1 | All history | 21 | 0.0135 | 0.350 |
| 10 | Running max | 3 | All history | 21 | 0.0135 | 0.350 |
| 10 | Running max | 6 | All history | 21 | 0.0135 | 0.350 |
| 10 | Mean score | 1 | 1 month | 24 | 0.0154 | 0.400 |
| 10 | Mean score | 3 | 3 months | 20 | 0.0128 | 0.333 |
| 10 | Mean score | 6 | 6 months | 23 | 0.0147 | 0.383 |
| 20 | Running max | 1 | All history | 34 | 0.0109 | 0.567 |
| 20 | Running max | 3 | All history | 34 | 0.0109 | 0.567 |
| 20 | Running max | 6 | All history | 34 | 0.0109 | 0.567 |
| 20 | Mean score | 1 | 1 month | 35 | 0.0112 | 0.583 |
| 20 | Mean score | 3 | 3 months | 37 | 0.0119 | 0.617 |
| 20 | Mean score | 6 | 6 months | 38 | 0.0122 | 0.633 |
| 50 | Running max | 1 | All history | 44 | 0.0056 | 0.733 |
| 50 | Running max | 3 | All history | 45 | 0.0058 | 0.750 |
| 50 | Running max | 6 | All history | 45 | 0.0058 | 0.750 |
| 50 | Mean score | 1 | 1 month | 44 | 0.0056 | 0.733 |
| 50 | Mean score | 3 | 3 months | 43 | 0.0055 | 0.717 |
| 50 | Mean score | 6 | 6 months | 42 | 0.0054 | 0.700 |
| 100 | Running max | 1 | All history | 48 | 0.0031 | 0.800 |
| 100 | Running max | 3 | All history | 48 | 0.0031 | 0.800 |
| 100 | Running max | 6 | All history | 48 | 0.0031 | 0.800 |
| 100 | Mean score | 1 | 1 month | 46 | 0.0029 | 0.767 |
| 100 | Mean score | 3 | 3 months | 45 | 0.0029 | 0.750 |
| 100 | Mean score | 6 | 6 months | 45 | 0.0029 | 0.750 |
| 200 | Running max | 1 | All history | 50 | 0.0016 | 0.833 |
| 200 | Running max | 3 | All history | 50 | 0.0016 | 0.833 |
| 200 | Running max | 6 | All history | 50 | 0.0016 | 0.833 |
| 200 | Mean score | 1 | 1 month | 47 | 0.0015 | 0.783 |
| 200 | Mean score | 3 | 3 months | 45 | 0.0014 | 0.750 |
| 200 | Mean score | 6 | 6 months | 45 | 0.0014 | 0.750 |

*Per-run capacity scaled with cadence so cumulative workload matched the nominal monthly budget. Under the running-maximum rule all 60 cases were eligible; shorter score windows reduced eligibility to 75-90%.*

#### Table S13. Validation of the GPT-5 colonoscopy annotation pipeline (n = 50)

| Task | Correct (n) | Reviewed (n) | Accuracy |
|---|---|---|---|
| Identification of colonoscopy reports | 50 | 50 | 1.00 |
| Detection of inflammation (present vs absent) | 48 | 50 | 0.96 |
| Classification of inflammation type | 45 | 50 | 0.90 |

#### Table S14. Subgroup performance at external validation (ZSFG)

| Subgroup | n | Cases | AUROC | AUPRC | Brier | TPR | PPV | PPR@thr |
|---|---|---|---|---|---|---|---|---|
| Sex: Female | 777 | 55 | 0.972 | 0.786 | 0.101 | 0.909 | 0.360 | 0.179 |
| Sex: Male | 757 | 84 | 0.983 | 0.894 | 0.112 | 0.953 | 0.435 | 0.245 |
| Sex: Nonbinary | 2 | 0 | NE | - | 0.000 | - | - | 0.000 |
| Race: American Indian/AK Native † | 17 | 2 | 1.000 | 1.000 | 0.117 | 1.000 | 0.500 | 0.235 |
| Race: Asian | 341 | 26 | 0.970 | 0.767 | 0.115 | 0.885 | 0.354 | 0.191 |
| Race: Black/African American | 256 | 24 | 0.968 | 0.888 | 0.121 | 0.880 | 0.379 | 0.226 |
| Race: Decline to answer | 35 | 0 | NE | - | 0.128 | - | - | 0.143 |
| Race: Native Hawaiian/Pac. Isl. † | 10 | 2 | 0.938 | 0.833 | 0.103 | 1.000 | 0.667 | 0.300 |
| Race: Other | 541 | 41 | 0.979 | 0.839 | 0.098 | 0.927 | 0.365 | 0.192 |
| Race: Unknown | 9 | 0 | NE | - | 0.104 | - | - | 0.222 |
| Race: White | 328 | 44 | 0.991 | 0.906 | 0.095 | 1.000 | 0.524 | 0.256 |
| Ethnicity: Decline to answer | 17 | 0 | NE | - | 0.057 | - | - | 0.059 |
| Ethnicity: Not Hispanic/Latino | 952 | 92 | 0.978 | 0.846 | 0.107 | 0.935 | 0.422 | 0.216 |
| Ethnicity: Hispanic/Latino | 567 | 47 | 0.980 | 0.856 | 0.106 | 0.936 | 0.373 | 0.208 |
| Age: 18-29 | 263 | 12 | 0.984 | 0.671 | 0.128 | 1.000 | 0.226 | 0.202 |
| Age: 30-49 | 540 | 50 | 0.972 | 0.845 | 0.134 | 0.922 | 0.359 | 0.242 |
| Age: 50+ | 734 | 77 | 0.984 | 0.923 | 0.078 | 0.935 | 0.511 | 0.192 |

*TPR = true-positive rate (sensitivity); PPV = positive predictive value; PPR@thr = positive-prediction rate at the operating threshold; Cases = incident cases in subgroup. Metrics computed at the fixed operating threshold.*

*NE = not evaluable (no incident cases in subgroup). † Fewer than 10 incident cases; estimates unstable and excluded from the AUROC range cited in the main text (0.968-0.991, adequately powered subgroups). Across adequately powered subgroups, discrimination, calibration (Brier), true-positive-rate, and positive-prediction-rate were consistent; across-group PPV differences tracked underlying case prevalence.*

#### Supplementary Figures

**Supplementary Figure S1. Calibration of the Mamba model before and after isotonic recalibration.** Predicted probability of incident IBD (x-axis) versus observed proportion (y-axis) in the internal UCSF test set, shown (A) before and (B) after isotonic recalibration fitted on the validation set. The diagonal indicates perfect calibration; points below it indicate overprediction and points above indicate underprediction. Curves reflect the 1:10 case-to-control sampling ratio and will shift at deployment prevalence. IBD, inflammatory bowel disease; UCSF, University of California, San Francisco.

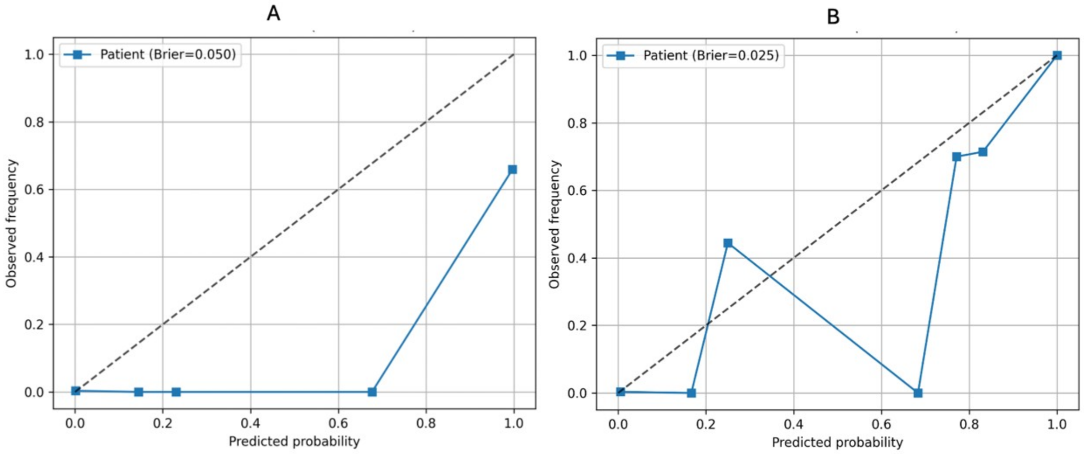


(A) Before isotonic regression. (B) After isotonic regression. Patient-level, internal test set.

**Supplementary Figure S2. Temporal robustness of model discrimination, 2012–2024.** Discrimination of the Mamba model in the internal test set by index year (year of diagnosis for cases and of last encounter for controls), grouped into four multi-year bins spanning 2012–2024. Bins contained 27 (7 cases), 31 (4 cases), 34 (8 cases), and 212 (11 cases) patients; estimates for bins with few cases are imprecise. Points show the area under the receiver operating characteristic curve (AUROC) and the area under the precision–recall curve (AUPRC) within each bin. Temporal drift was assessed by logistic regression of case status on calendar year, prediction score, and their interaction; a non-significant year-by-score interaction indicates that discrimination did not change materially over the study period. AUPRC, area under the precision–recall curve; AUROC, area under the receiver operating characteristic curve; CI, confidence interval.

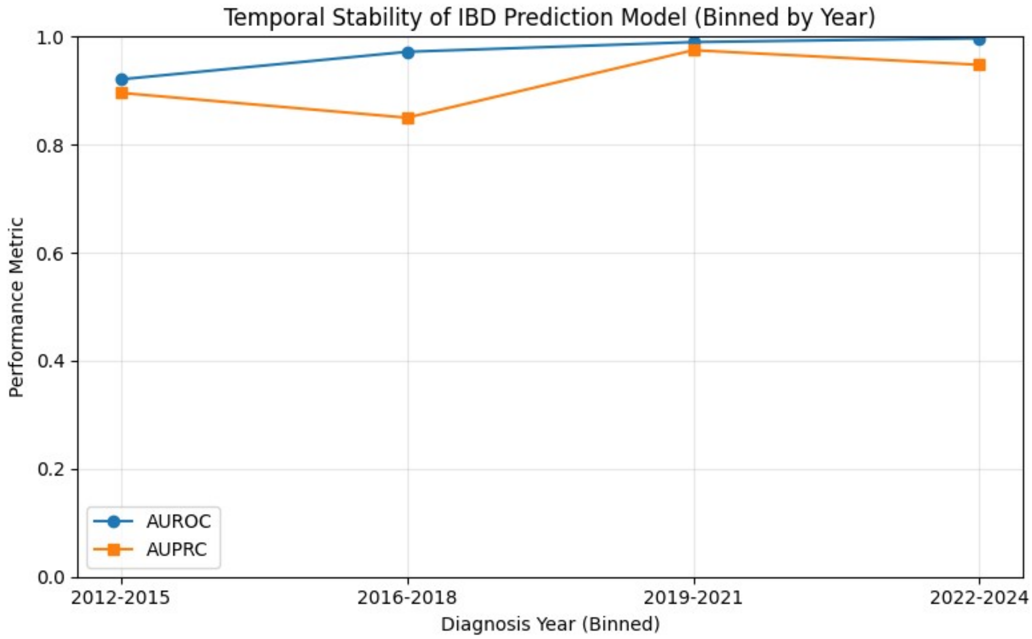
